# Investigating the Causal Relationship between Neuroticism and Alzheimer’s Disease Using Mendelian Randomization and the Mediating Role of Modifiable Risk Factors

**DOI:** 10.64898/2026.08.01.26359481

**Authors:** Niki Akbarian, Mahbod Ebrahimi, Fernanda C. Dos Santos, Sara Sadat Afjeh, Mohamed Abdelhack, Andreea O. Diaconescu, Daniel Felsky, Clement C. Zai, James L Kennedy

**Affiliations:** Tanenbaum Centre for Pharmacogenetics, Campbell Family Mental Health Research Institute, Centre for Addiction and Mental Health, Toronto, Ontario, Canada; Dept of Psychiatry and Institute of Medical Science, University of Toronto, Ontario, Canada; Krembil Centre for Neuroinformatics, Centre for Addiction and Mental Health, Toronto, Ontario, Canada; Dalla Lana School of Public Health, University of Toronto, Ontario, Canada; Department of Laboratory Medicine and Pathobiology, University of Toronto, Ontario, Canada

**Keywords:** Neuroticism, Alzheimer’s Disease, Mendelian Randomization, Depression, Hypertension, Alcohol Use, Mediation

## Abstract

Neuroticism, a personality trait characterized by the predisposition to experience intense and frequent negative emotions, has been associated with an increased risk of Alzheimer’s disease (AD). However, the mechanisms underlying this association remain unclear. Our study investigated two potential pathways: (1) whether the relationship between neuroticism and AD is causal, and (2) whether it is mediated by health and behavioral factors associated with both neuroticism and AD risk. To assess causality, a two-sample Mendelian randomization (MR) was employed using publicly available genome-wide association studies (GWAS) for neuroticism (Nagel et al., 2018) and AD (Bellenguez et al., 2022). Mediation analysis was conducted in a subset of UK Biobank participants aged 60 and older, including 121,825 controls (mean age = 63.9 ± 2.81; 61,993 females or 50.9%) and 1,277 individuals with AD (mean age = 65.6 ± 2.71; 628 females or 49.2%). All participants had complete data on neuroticism and the potential mediators. MR analysis suggested that the neuroticism-AD relationship is unlikely to be causal. However, depression (β=0.048, p=3×10⁻⁴), hypertension (β=0.005, p=2×10⁻⁴), and alcohol consumption (β=0.001, p=1×10⁻^5^) were identified as significant mediators of the relationship between higher neuroticism and increased AD risk. Overall, the association between neuroticism and AD may be largely explained by modifiable health and behavioral factors rather than a direct causal effect. A better understanding of these mediating pathways may inform targeted prevention and therapeutic strategies to reduce AD risk.

## 1. Introduction

Alzheimer’s Disease (AD) is a progressive neurodegenerative disease that is the most common cause of dementia (Kumar et al., 2024). Each year, more than 55 million individuals are affected by dementia worldwide, and AD contributes to up to 70% of the cases (Shin, 2022). Since the pathological changes associated with AD are progressive and are present years to decades before the symptoms are apparent (Bateman et al., 2012; Sperling et al., 2011), it is crucial to identify at-risk individuals as early as possible to implement potential preventive interventions.

The personality trait of neuroticism, characterized by the predisposition to experience intense and frequent negative emotions (Barlow et al., 2014), has been identified by longitudinal epidemiological studies as a factor that increases the risk of developing AD (Aschwanden et al., 2021; Beck et al., 2024; D’Iorio et al., 2018; Johansson et al., 2014; Terracciano et al., 2021). However, the underlying mechanisms of this association remain unknown.

Twin studies estimate the heritability of neuroticism to range between 30% and 60%, highlighting a substantial genetic contribution to this personality trait (Hettema et al., 2004). Neuroticism is also recognized as a polygenic trait, with Genome-Wide Association Studies (GWAS) identifying multiple genetic loci associated with it (Nagel et al., 2018). These neuroticism-associated genes are primarily expressed in brain regions such as the anterior cingulate cortex, cerebellar hemisphere, frontal cortex, and nucleus accumbens of the basal ganglia (Nagel et al., 2018). Although not reaching statistical significance, expression of these genes has also been observed in the amygdala, caudate nucleus, hippocampus, and hypothalamus. In addition, neuroticism has been associated with seven gene ontology (GO) sets, most notably those related to neurogenesis and neuronal differentiation (Nagel et al., 2018). The genetic, transcriptomic, and neuroanatomical overlap between neuroticism and AD pathology suggests a potential biological pathway through which neuroticism could influence AD risk. This raises the hypothesis that neuroticism may causally influence AD risk. However, establishing causality requires methods beyond observational or correlational, such as Mendelian randomization (MR). A limited number of studies have previously applied MR to examine the relationship between neuroticism and AD, including Ma et al. (2021) and Zang et al. (2021). While these studies relied on earlier GWAS datasets and different MR frameworks (e.g., GSMR), the present study leverages more recent and larger GWAS for both neuroticism and AD and incorporates additional sensitivity analyses, including MR-PRESSO, to assess the robustness of MR assumptions, providing a complementary and updated evaluation of the potential causal relationship between neuroticism and AD.

In parallel, neuroticism has been associated with a range of health and behavioral factors, including hypertension (Lone and Othman Albotuaiba, 2023; Zhang et al., 2021), depression (Kotov et al., 2010; Speed et al., 2019), lower educational attainment (Chamorro-Premuzic and Furnham, 2003; Mõttus et al., 2017), smoking (Hakulinen et al., 2015; Zvolensky et al., 2015), alcohol use (Adan et al., 2017; Turiano et al., 2012), and abnormal body weight (Ferguson et al., 2012; Sutin and Terracciano, 2016). These factors are themselves recognized as modifiable risk factors for AD (Livingston et al., 2020). Together, these findings suggest that health and behavioral factors may partially account for the observed association between neuroticism and AD.

Emerging research has begun to examine factors that may mediate the association between neuroticism and Alzheimer’s disease (AD). For example, Duchek et al. (2020) examined whether AD-related biomarkers, including amyloid burden, cerebrospinal fluid Aβ42 and tau levels, and hippocampal volume, were associated with the relationship between neuroticism and Alzheimer’s dementia diagnosis. Similarly, Terracciano et al. (2022) reported associations between higher neuroticism and greater amyloid and tau pathology, while Montoliu et al. (2022) identified perceived stress as a factor statistically linking neuroticism to cognitive decline. Additional studies have implicated structural brain changes and lifestyle-related factors, such as reduced gray matter volume, increased white matter hyperintensities, and lower engagement in physical and cognitive activities (Gao et al., 2024; Stephan et al., 2024; Terracciano et al., 2023). Despite these findings, no study to date has specifically examined whether modifiable health and behavioral factors statistically account for the association between neuroticism and AD.

As a result, the overall aim of this study was twofold; first, to search for causality between neuroticism and AD using Mendelian randomization, and second, to investigate whether this relationship is mediated by modifiable health and behavioral factors.

## 2. Methods

### 2.1 Study Population

Participants in this study were selected from the UK Biobank (Sudlow et al., 2015), a large prospective cohort study. Between 2006 and 2010, the UK Biobank recruited 502,633 participants aged 40 to 69 years from 22 assessment centres across Scotland, England, and Wales. The study collected extensive data on family history, early life exposures, lifestyle and psychosocial factors, as well as ICD-coded diagnoses, referrals, and prescriptions derived from participants’ health records.

For the present analysis, inclusion criteria consisted of individuals aged 60 years or older with complete data on the Eysenck Personality Questionnaire-Revised Short Form (Eysenck et al., 1991), which measures neuroticism, and on all potential mediators investigated in the study: educational attainment, body mass index (BMI), smoking status, alcohol intake frequency, and history of hypertension and depressive episodes.

### 2.2 Measures and Diagnoses

#### 2.2.1 Alzheimer’s Disease Diagnosis

The sample included individuals with AD and controls who were free of AD. The diagnosis of participants with AD was determined using the ICD-10 codes G30 (Data-Field 131036) and F00 (Data-Field 130836 and Data-Field 42020). In total, 1,277 participants were diagnosed with AD and 121,825 participants were considered as controls. Information on the timing of diagnoses was not incorporated, and case status was treated without modeling time of occurrence.

#### 2.2.2 Neuroticism

The 12-item Neuroticism subscale of the Eysenck Personality Questionnaire-Revised Short Form (EPQ-R) was administered to assess neuroticism (Eysenck et al., 1991). In the EPQ-R, traits such as loneliness, nervousness, irritability, mood swings, guilt, feeling fed up, being a worrier, and experiencing hurt feelings were assessed. Response options were as follows: “Yes” (coded as 1), “No” (coded as 0), and “Do not know” or “Prefer not to answer” (both coded as missing). Consequently, a participant could attain a maximum score of 12 on the neuroticism scale by answering "Yes" to all questions. Neuroticism scores were standardized prior to analyses.

In the total sample of the UK Biobank (N=502,413), 100,914 participants were missing data on the neuroticism score, resulting in a missingness rate of 20.09%. For this study, only participants with available neuroticism scores were included.

#### 2.2.3 Educational Attainment

Participants were asked to report their highest qualification achieved (Data-Field 6138) from a list of options, including college or university degree, Advanced/Advanced Subsidiary (A/AS) levels, Ordinary (O) levels/ General Certificate of Secondary Educations (GCSEs), Certificate of Secondary Educations (CSE), National Vocational Qualifications (NVQ), Higher National Diploma (HND)/Higher National Certificate (HNC) or equivalent, other professional qualifications (e.g., nursing, teaching), none of the above, or prefer not to answer (Data-Field 6138). The qualifications were then converted based on the International Standard Classification for Education (ISCED) coding for years of education, as used in previous studies from the UK Biobank (Carter et al., 2022; Lee et al., 2018). That is, college or university degree, A/AS, O/GCSEs, CSEs, NVQ/HND/HNC, and other qualifications were coded as 20 years, 13 years, 10 years, 10 years, 19 years, and 15 years, respectively. Subsequently, years of education were treated as a continuous variable for the statistical analysis.

Out of all participants in the UK Biobank, 95,391 individuals did not have data available for their educational attainment, resulting in a missing rate of 19.00%. These individuals were excluded from this study.

#### 2.2.4 Body Mass Index

During their visit to the assessment centre, the body mass index (BMI) of participants (Data-Field 21001) was utilized as a proxy for obesity. All the participants had data on BMI.

#### 2.2.5 Smoking Status

Participants were queried regarding their smoking habits, and they were categorized into three groups based on their smoking status; current smokers, past smokers, or those who have never smoked (Data-Field 20116). 2,057 participants responded “prefer not to answer” to the question regarding their smoking habits and 893 participants did not respond to the questions at all. All these participants were excluded from the analysis. Consequently, smoking status was coded as a binary variable, with ever smokers (current or previous) coded as 1 and never smokers as 0.

#### 2.2.6 Alcohol Intake Frequency

Participants were surveyed on their average alcohol intake (Data-Field 1558) with response options including daily or almost daily (coded as 5), three or four times a week (coded as 4), once or twice a week (coded as 3), once or three times a month (coded as 2), special occasions only (coded as 1), and never (coded as 0). Ultimately, alcohol intake frequency was treated as a continuous variable. Of 502,413 participants, 603 chose not to disclose their answers regarding their alcohol intake frequency and 899 did not respond to the questionnaire, resulting in a missing rate of 0.29%. These individuals were excluded from the study.

#### 2.2.7 History of Hypertension

Hypertension status was determined by ICD-10 code I10 for essential hypertension in the UK Biobank (Data-Field 41270). Individuals with a lifetime history of hypertension were assigned a code 1 (39,605 of 502,413 participants), while those without (92.12% of participants) were assigned a code of 0.

#### 2.2.8 History of Depressive Disorders

To identify participants with a lifetime history of depressive episodes, ICD-10 code F32 from Data-Field 41270 was used. Individuals with a documented lifetime history of depressive episodes were assigned a code of 1 (4,468 of 502,413 participants), while those without (99.11% of participants) were assigned a code of 0.

### 2.3 Statistical Analysis

#### 2.3.1 Mendelian Randomization (MR)

MR is a genetic technique that uses single-nucleotide polymorphisms (SNPs) to mimic the design of randomized controlled trials. It is based on Mendel’s laws of inheritance and the fixed nature of germline genotypes (Richmond and Davey Smith, 2022). In MR, SNPs function as instrumental variables, and observing an individual’s genotype at a particular SNP is akin to the random assignment of an individual to either an exposed group or a control group in a randomized controlled trial (Davey Smith and Hemani, 2014). There are different approaches to MR. The most common approach is a 2-sample MR, where the SNP-exposure effects and the SNP-outcome effects are obtained from two separate studies (Burgess et al., 2019).

For this study, we used the MR-Base platform (Hemani et al., 2018) to run 2-sample MR, which uses Wald ratios of SNPs to assess the causal relationship between the exposure and outcome. To perform 2-sample MR, we first extracted significant SNPs (p<5×10^−8^) from the most recent summary-level GWAS on neuroticism by Nagel et al. (2018). Subsequently, the same SNPs were extracted from the summary-level GWAS of AD by Bellenguez et al. (2022). Next, Inverse Variance Weighting (IVW) was used to conduct MR.

Since IVW assumes that there is no horizontal pleiotropy in the data, meaning that the SNPs affect the outcome only through the exposure and not through other pathways (Boehm and Zhou, 2022), additional MR models robust to horizontal pleiotropy, including Egger regression, weighted median, and weighted mode-based, were used to validate the results from IVW.

Moreover, because the GWAS datasets for neuroticism and AD both included participants from the UK Biobank, raising concerns about sample overlap, the analysis was repeated using an alternative AD GWAS by Kunkle et al. (2019), which excluded UK Biobank participants. In addition, leave-one-out analysis, Cochran’s Q test, and MR-Egger intercept were conducted as sensitivity analyses to detect heterogeneity and potential pleiotropy among SNPs.

#### 2.3.2 Mediation Analysis

The total effect of neuroticism scores on the probability of AD was first assessed using logistic regression (Baron and Kenny, 1986). In the model, AD diagnosis served as the dependent variable, neuroticism as the independent variable, and age and sex as covariates. After confirming that the total effect of neuroticism on AD diagnosis was statistically significant, structural equation modeling (SEM) was used to evaluate the mediating effect of the potential mediators in the relationship between neuroticism and the probability of AD. For the mediation analysis using SEM, the direct and indirect effects were examined. The direct effect represents the effect of an independent variable (neuroticism) on the outcome (probability of AD) that is not mediated by any of the mediators. The indirect effect refers to the pathway from the independent variable to the outcome through the mediators. Last, the total effect is the sum of the direct and indirect effects of the independent variable on the outcome. We estimated a multivariate mediation model including all candidate mediators simultaneously to account for intercorrelations among mediators and avoid inflated indirect effects. As a sensitivity analysis, we also ran separate mediation models for each mediator to confirm the robustness of the indirect effects. All the analyses were performed by RStudio v4.3.3 software (RStudio, 2024). Specifically, the mediation analysis was conducted with the “sem” function of the “lavaan” package in R (Rosseel, 2012). Furthermore, age and sex were included as covariates in all regression and mediation models. To calculate 95% confidence intervals (CI), bootstrapping with 1000 bootstrap samples was used. Moreover, the false discovery rate (FDR) method was used to adjust p-values for multiple comparisons.

### 2.4 Ethical Approval

The UK Biobank received ethics approval from the Northwest Multi-Centre Research Ethics Committee. All participants provided written consent to the study, and any participant who withdrew from the study was removed from our analysis. The data was accessed under the application #61530 with the Centre for Addiction and Mental Health as the lead institution.

## 3. Results

### 3.1 Demographics

The demographic characteristics of participants are summarized in Table 1. In total, 123,102 participants (62,621 females) with a mean age of 63.9 (SD=2.81) were included in the study.

**Table 1.** Demographic Characteristics of Participants. N=Sample Size, SD=Standard Deviation.

|  | **AD Cases**<br>**(N=1,277)** | **Controls**<br>**(N=121,825)** | **Overall**<br>**(N=123,102)** |
| --- | --- | --- | --- |
| **Sex** |  |  |  |
| Female | 628 (49.2%) | 61993 (50.9%) | 62621 (50.9%) |
| Male | 649 (50.8%) | 59832 (49.1%) | 60481 (49.1%) |
| **Age** |  |  |  |
| Mean (SD) | 65.6 (2.71) | 63.9 (2.81) | 63.9 (2.81) |
| **Neuroticism** |  |  |  |
| Mean (SD) | 3.64 (3.12) | 3.59 (3.06) | 3.59 (3.06) |
| **Educational Attainment (Years)** |  |  |  |
| Mean (SD) | 15.1 (4.48) | 15.3 (4.49) | 15.3 (4.49) |
| **BMI** |  |  |  |
| Mean (SD) | 27.1 (4.68) | 27.3 (4.44) | 27.3 (4.44) |
| **Smoking Status** |  |  |  |
| Current | 88 (6.9%) | 8595 (7.1%) | 8683 (7.1%) |
| Never | 639 (50.0%) | 62811 (51.6%) | 63450 (51.5%) |
| Previous | 550 (43.1%) | 50419 (41.4%) | 50969 (41.4%) |
| **Alcohol Intake Frequency** |  |  |  |
| Never | 146 (11.4%) | 8468 (7.0%) | 8614 (7.0%) |
| Special occasions only | 148 (11.6%) | 12459 (10.2%) | 12607 (10.2%) |
| One to three times a month | 123 (9.6%) | 11584 (9.5%) | 11707 (9.5%) |

|  | **AD Cases**<br>**(N=1,277)** | **Controls**<br>**(N=121,825)** | **Overall**<br>**(N=123,102)** |
| --- | --- | --- | --- |
| Once or twice a week | 292 (22.9%) | 27336 (22.4%) | 27628 (22.4%) |
| Three or four times a week | 268 (21.0%) | 28832 (23.7%) | 29100 (23.6%) |
| Daily or almost daily | 300 (23.5%) | 33146 (27.2%) | 33446 (27.2%) |
| **Hypertension** |  |  |  |
| No | 1080 (84.6%) | 108773 (89.3%) | 109853 (89.2%) |
| Yes | 197 (15.4%) | 13052 (10.7%) | 13249 (10.8%) |
| **Depression** |  |  |  |
| No | 1256 (98.4%) | 121064 (99.4%) | 122320 (99.4%) |
| Yes | 21 (1.6%) | 761 (0.6%) | 782 (0.6%) |

Of those, 1,277 (mean age=65.6 (SD=2.71); 628 females or 49.2%) were diagnosed with AD, and the rest were controls (N=121,825; mean age=63.9 (SD=2.81); 61,993 females or 50.9%). In the overall sample, the mean neuroticism score was 3.59 (SD=3.06), 13,249 (10.8%) participants had hypertension, and 782 (0.6%) participants had depression. The details of other characteristics of each group are demonstrated in Table 1.

### 3.2 MR Findings on the Neuroticism–Alzheimer’s Disease Relationship

All MR models indicated a weak positive association between SNPs associated with neuroticism and AD, as reflected in the positive slopes shown in Figure 1. As shown in Table 2, the IVW model yielded a statistically significant estimate (β=0.0008, SE=0.0002, *p*=0.001), as did the weighted median model (β=0.0007, SE=0.0003, *p*=0.012), although the effect sizes were small. In contrast, results from the MR-Egger regression (β=0.0010, SE=0.00018, *p*=0.578) and weighted mode model (β=0.0006, SE=0.0005, *p*=0.286) were not statistically significant.

**Figure 1.**
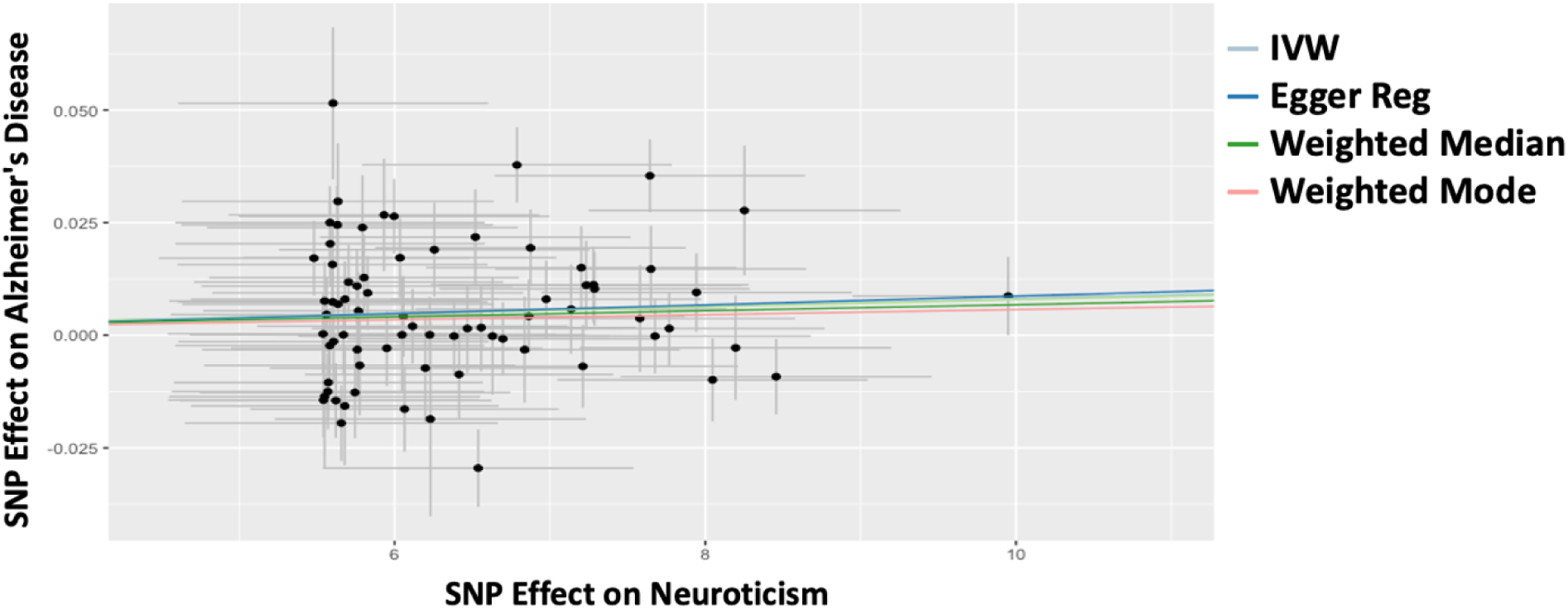
Scatterplot of SNP Effects on Neuroticism (x-axis) and Alzheimer’s Disease (y-axis), with Mendelian Randomization (MR) Estimates. Each point represents a SNP, with horizontal and vertical lines indicating standard errors for the exposure (neuroticism) and outcome (AD), respectively. The overlaid lines represent causal effect estimates derived from four MR methods: inverse-variance weighted (IVW, light blue), MR-Egger regression (Egger Reg, dark blue), weighted median (green), and weighted mode (pink).

**Table 2.** Results of the MR Analysis Using Different Models Investigating the Causal Relationship between Neuroticism and Alzheimer’s Disease. . N SNP=Number of SNPs used for each model, SE=Standard Error.

| Method | N SNP | Estimate | SE | p-value |
| --- | --- | --- | --- | --- |
| Inverse Variance Weighted | 74 | 0.0008 | 0.0002 | **0.001** |
| Egger Regression | 74 | 0.0010 | 0.0018 | 0.578 |
| Weighted Median | 74 | 0.0007 | 0.0003 | **0.012** |
| Weighted Mode | 74 | 0.0006 | 0.0005 | 0.286 |

The MR-Egger intercept was -0.0011 (SE=0.0112, *p*=0.9253). This non-significant and close to zero intercept suggested no evidence of directional pleiotropy or unbalanced heterogeneity among SNPs.

The Cochran’s Q test was performed for both the IVW (Q=163.007, df=73, *p*=7.96×10⁻⁹) and MR-Egger (Q=162.99, df=72, *p*=5.25×10⁻⁹) models, indicating significant heterogeneity among SNPs, suggesting that not all SNPs were valid instruments fulfilling the assumptions of these MR models. Therefore, a leave-one-out analysis as well as MR-Radial and MR-PRESSO were conducted to evaluate the influence of individual SNPs on the overall results, identify outlier SNPs, and reassess the causal relationship between neuroticism and AD after excluding them.

As shown in Figure 2, which depicts the leave-one-out analysis, the causal estimate of neuroticism on AD remained stable regardless of which SNP was excluded, proposing that no single SNP disproportionately influenced the results. Nevertheless, MR-Radial and MR-PRESSO identified three potential outlier SNPs (i.e., rs11509880, rs3751855, and rs7107356). The estimate of the IVW model remained significant, yet very small, after excluding potential outlier SNPs (β=0.0007, SE=0.0002, *p*=0.002), depicted in Figure 3.

**Figure 2.**
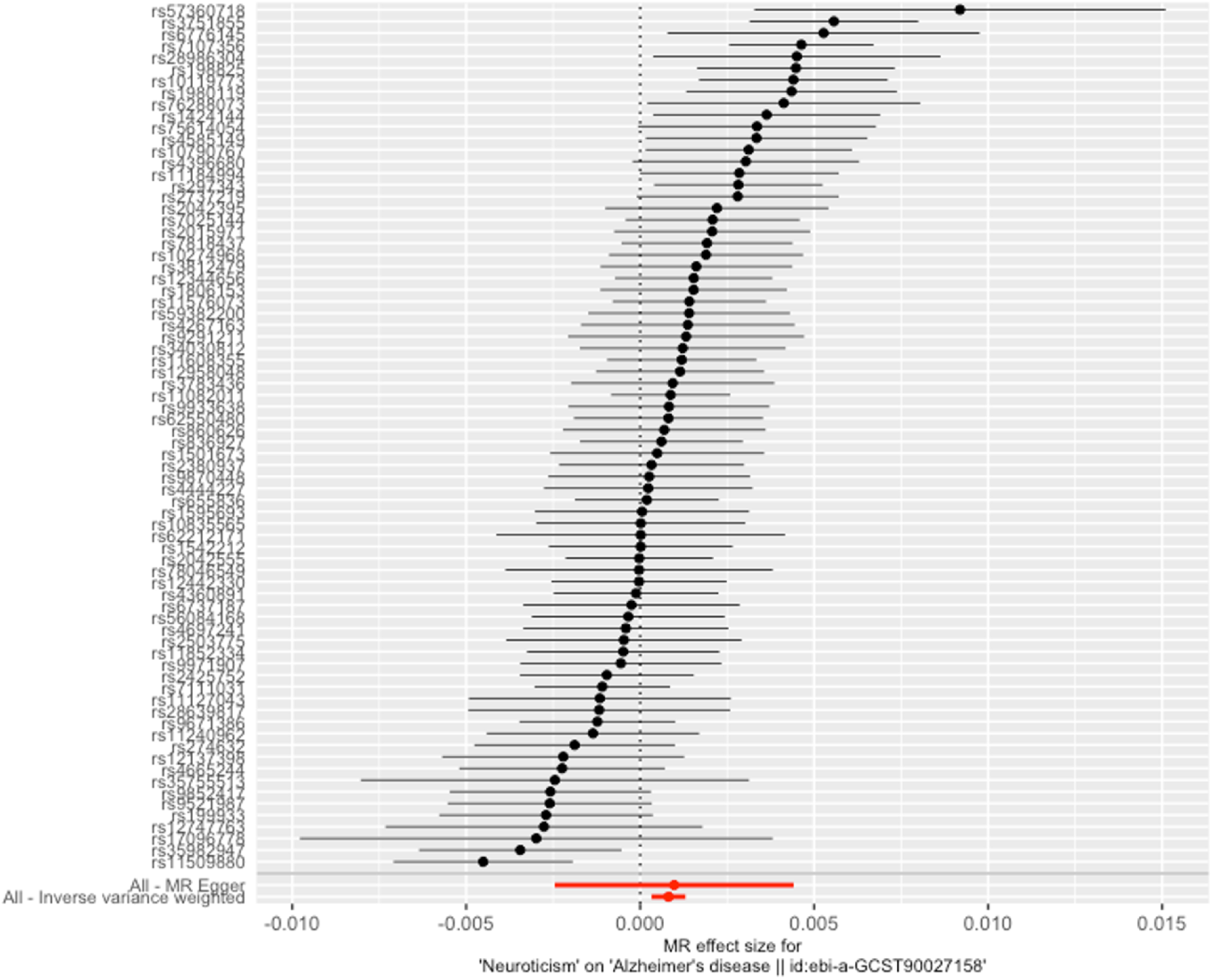
Leave-One-Out Sensitivity Analysis of the Effect of Neuroticism on Alzheimer’s Disease. Each black point represents the Mendelian Randomization (MR) estimate obtained by excluding one single nucleotide polymorphisms (SNPs) at a time. The corresponding identifiers for the SNPs excluded are indicated on the left (e.g., rs#). The Horizontal lines represent 95% confidence intervals of MR estimates. At the bottom, the overall MR estimates with all SNPs included using MR-Egger regression (top red dot) and inverse-variance weighting (bottom red dot) are shown.

**Figure 3.**
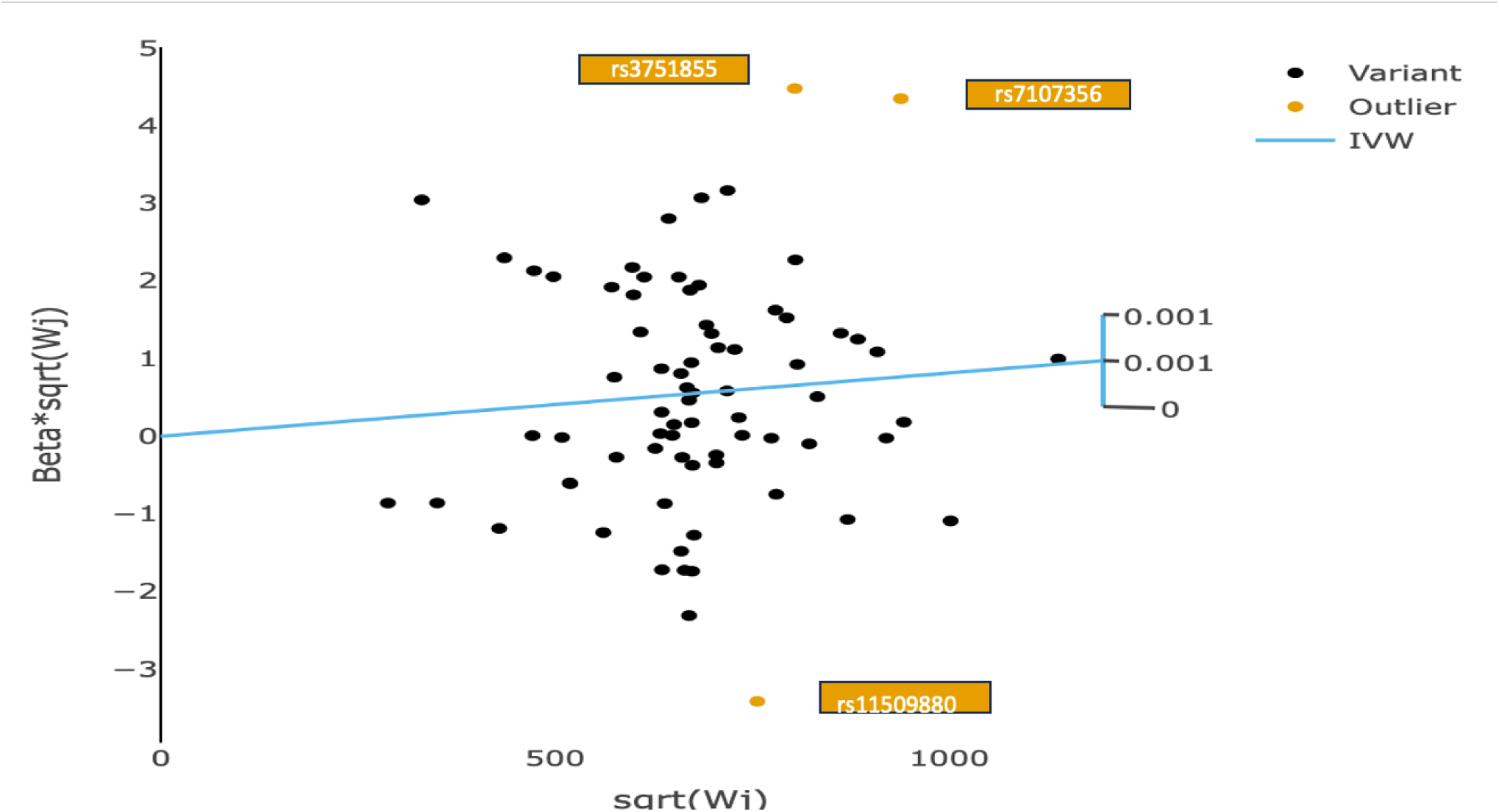
Radial Plot Detecting Outlier SNPs in Mendelian Randomization Analysis. The plot visualizes the contribution of each single nucleotide polymorphism (SNP), represented as black dots, to the overall Mendelian Randomization (MR) estimate. The x-axis represents the square root of the inverse variance of each SNP-outcome association and the y-axis shows the MR estimate for each SNP weighted by its precision. The blue line represents the inverse-variance model and the yellow dots are detected outlier SNPs.

When the analysis was repeated using an AD GWAS that did not include UK Biobank participants, the direction of the association between SNPs associated with neuroticism and AD remained positive (Figure 4). However, as shown in Table 3, the results were no longer statistically significant across any of the MR models. In addition, the MR-Egger intercept was -0.0180 (SE=0.0163, *p*=0.2727), indicating no evidence of directional pleiotropy or unbalanced heterogeneity. However, Cochran’s test for both the IVW (Q=103.213, df=70, *p*=0.0060) and MR-Egger (Q=101.41, df=69, *p*=0.0067) suggested significant heterogeneity among the SNPs.

**Figure 4.**
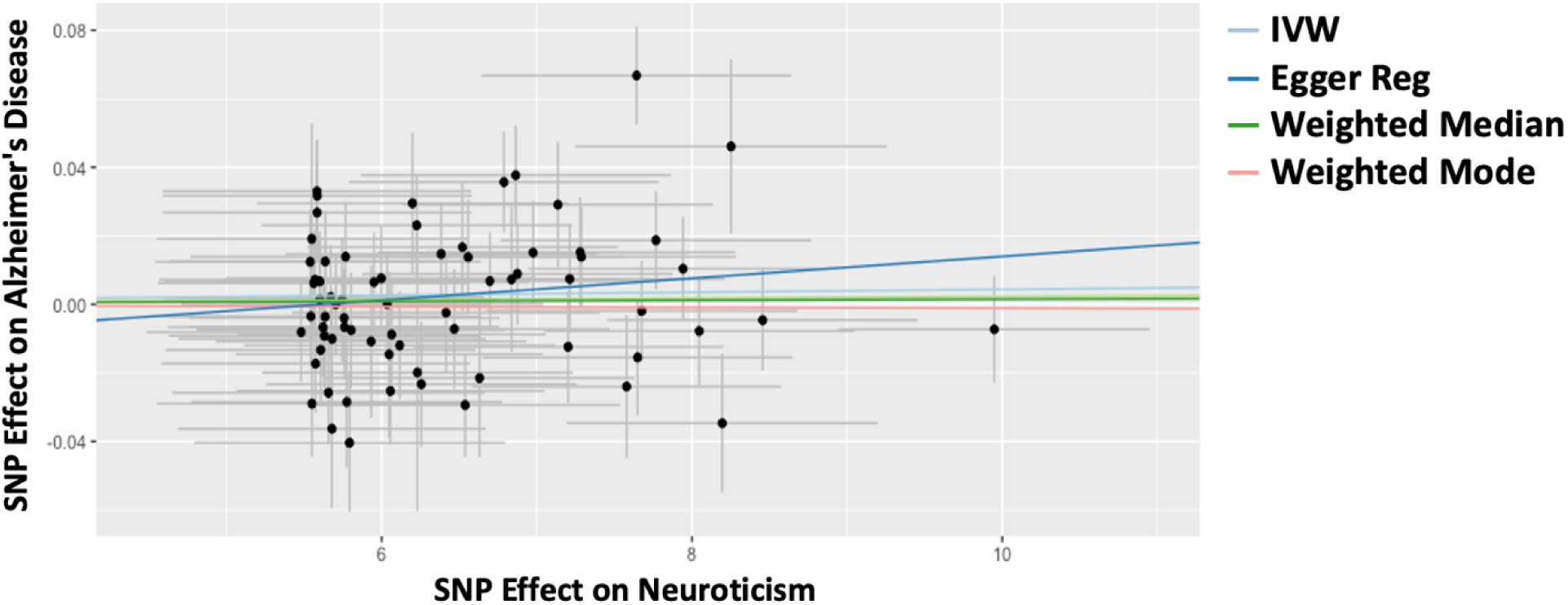
Scatterplot of SNP Effects on Neuroticism (x-axis) and Alzheimer’s Disease (y-axis) Based on Mendelian Randomization Using GWAS with Non-Overlapping Samples. Each point represents a SNP, with horizontal and vertical lines indicating standard errors for the exposure (neuroticism) and outcome (AD), respectively. The overlaid lines represent causal effect estimates derived from four MR methods: inverse-variance weighted (IVW, light blue), MR-Egger regression (Egger Reg, dark blue), weighted median (green), and weighted mode (pink).

**Table 3.** Results of the MR Models Investigating the Causal Relationship between Neuroticism and Alzheimer’s Disease Using GWAS with Non-Overlapping Samples. . N SNP=Number of SNPs used for each model, SE=Standard Error.

| Method | N SNP | Estimate | SE | p-value |
| --- | --- | --- | --- | --- |
| Inverse Variance Weighted | 71 | 0.0004 | 0.0003 | 0.223 |
| Egger Regression | 71 | 0.0032 | 0.0025 | 0.208 |
| Weighted Median | 71 | 0.0002 | 0.0005 | 0.744 |
| Weighted Mode | 71 | -0.0001 | 0.0009 | 0.905 |

**Table 4.** Logistic Regression Model Examining the Relationship Between Neuroticism (Z-Scores) and the Likelihood of AD Diagnosis. OR=Odds Ratio; 95% CI=95% Confidence Interval; SE=Standard Error; AIC=Akaike Information Criterion.

| Characteristic | Estimate | OR | 95% CI | SE | z-value | p-value |
| --- | --- | --- | --- | --- | --- | --- |
| **Intercept** | -18.227 | - | - | 0.661 | -27.568 | **<2 ×10<sup>-16</sup>** |
| **Neuroticism**<br>(Standardized) | 0.063 | 1.065 | 1.007, 1.125 | 0.028 | 2.231 | **0.026** |
| **Age** | 0.211 | 1.234 | 1.211, 1.259 | 0.010 | 20.917 | **2 ×10<sup>-16</sup>** |
| **Sex (Male)** | 0.052 | 1.053 | 0.942, 1.177 | 0.057 | 0.909 | 0.363 |

**AIC:** 22263

**Table 5.** Results of the Mediation Analysis with 1000 Bootstrapping Iterations. 95% CI= 95% Confidence Interval, SE= Standard Error. Adjusted p refers to p-values adjusted for multiple comparisons using the false discovery rate method. R^2^ explains the amount of variance of the likelihood of diagnosis with AD explained by each model. N represents standardized neuroticism scores. The model included age and sex as covariates.

|  | Estimate | 95% CI | SE | z-value | p-value | Adjusted p<br>(FDR) | R<sup>2</sup> |
| --- | --- | --- | --- | --- | --- | --- | --- |
| **Direct Effect** |  |  |  |  |  |  |  |
| N → AD | -0.146 | -0.315, -0.029 | 0.054 | -2.693 | **0.007** | – | – |
| **Indirect Effects** |  |  |  |  |  |  |  |
| N → **Hypertension** → AD | 0.005 | 0.003, 0.008 | 0.001 | 3.701 | **2×10<sup>-4</sup>** | **6×10<sup>-4</sup>** | 0.026 |
| N → **Smoking** → AD | 0.065 | 0.059, 0.262 | 0.045 | 1.383 | 0.167 | 0.210 | 0.024 |
| N → **BMI** → AD | 0.005 | -0.006, 0.011 | 0.004 | 1.252 | 0.210 | 0.210 | 0.005 |
| N → **Education** → AD | 0.046 | -0.050, 0.051 | 0.035 | 1.316 | 0.188 | 0.210 | 0.022 |
| N → **Alcohol** → AD | 0.001 | 9×10<sup>-4</sup>, 0.0021 | 3×10<sup>-4</sup> | 4.389 | **1×10<sup>-5</sup>** | **6×10<sup>-5</sup>** | 0.043 |
| N → **Depression** → AD | 0.048 | 0.017, 0.072 | 0.0134 | 3.593 | **3×10<sup>-4</sup>** | **7×10<sup>-4</sup>** | 0.089 |
| **Total Effect** |  |  |  |  |  |  |  |
| Neuroticism + Significant Mediators | 0.024 | 0.008, 0.049 | 0.010 | 2.311 | **0.021** | – | – |

### 3.3 Association Between Neuroticism and Alzheimer’s Disease Diagnosis, Unadjusted for the Potential Mediators: Logistic Regression Results

There was a statistically significant association between neuroticism (standardized) and the likelihood of an AD diagnosis (OR=1.065, 95% CI=[1.007, 1.125], *p*=0.026), as shown in Table 3, without accounting for the influence of potential mediators.

### 3.4 Identifying the Mediators of the Association between Neuroticism and AD

Hypertension (β=0.005, 95% CI=[0.003, 0.008], p=2.1×10⁻⁴), depression (β=0.048, 95% CI=[0.017, 0.072], p=3.3×10⁻⁴), and alcohol consumption (β=0.001, 95% CI=[9×10⁻⁴, 0.0021], p=1×10⁻⁵) were identified as significant mediators of the relationship between neuroticism and AD in the full SEM mediation model. Smoking status, BMI, and educational attainment were not significant mediators. Depression showed the strongest indirect effect compared to hypertension and alcohol intake frequency. The indirect effects through hypertension, depression, and alcohol consumption remained statistically significant after correction for multiple testing using FDR. To assess robustness, separate mediation models were run for each mediator individually. The pattern of results was consistent with the multivariate model as hypertension, alcohol intake, and depression showed significant indirect effects, whereas smoking, BMI, and education did not (Table 6).

**Table 6.** Separate Mediation Analyses Examining the Indirect Effects of Neuroticism on Alzheimer’s disease (AD) through Six Individual Potential Mediators. Each model included age and sex as covariates. For each mediator, the table reports the standardized regression coefficients (β) and p-values for the paths from neuroticism to the mediator and from the mediator to AD, as well as the indirect effect (ab), total effect (c + ab), and the significance of mediation. Significant indirect effects indicate evidence of mediation. Total effects reflect the overall association between neuroticism and AD in each model, regardless of whether mediation is significant.

| **Mediator** | **Neuroticism → Mediator**<br>**($\beta$, p)** | **Mediator → AD**<br>**($\beta$, p)** | **Indirect effect**<br>**($a*b$, p)** | **Total effect**<br>**($c + a*b$, p)** |
| --- | --- | --- | --- | --- |
| **Hypertension** | 0.015, $p < 0.001$ | 0.004, $p = 0.002$ | **$6.0 \times 10^{-5}$, $p = 0.002$** | 0.001, $p = 0.031$ |
| **Smoking** | 0.032, $p < 0.001$ | $< 0.001$, $p = 0.743$ | $< 0.001$, $p = 0.742$ | 0.001, $p = 0.034$ |
| **BMI** | -0.020, $p = 0.107$ | $< 0.001$, $p = 0.050$ | $< 0.001$, $p = 0.243$ | 0.001, $p = 0.039$ |
| **Education** | -0.207, $p < 0.001$ | $< 0.001$, $p = 0.197$ | $< 0.001$, $p = 0.198$ | 0.001, $p = 0.03$ |
| **Alcohol** | -0.039, $p < 0.001$ | -0.001, $p < 0.001$ | **$3.90 \times 10^{-5}$, $p < 0.001$** | 0.001, $p = 0.035$ |
| **Depression** | 0.006, $p < 0.001$ | 0.018, $p = 0.003$ | **$1.08 \times 10^{-4}$, $p = 0.003$** | 0.001, $p = 0.028$ |

Notably, in a mediation model for AD and neuroticism accounting for significant mediators, the direct association between neuroticism and AD remained significant, as illustrated in Figure 5. However, the association reversed in direction, becoming negative (β=-0.146, 95% CI=[-0.315, -0.029], *p*=0.007). In other words, after adjusting for hypertension, depression, and alcohol consumption, higher neuroticism scores were associated with a lower likelihood of developing AD.

**Figure 5.**
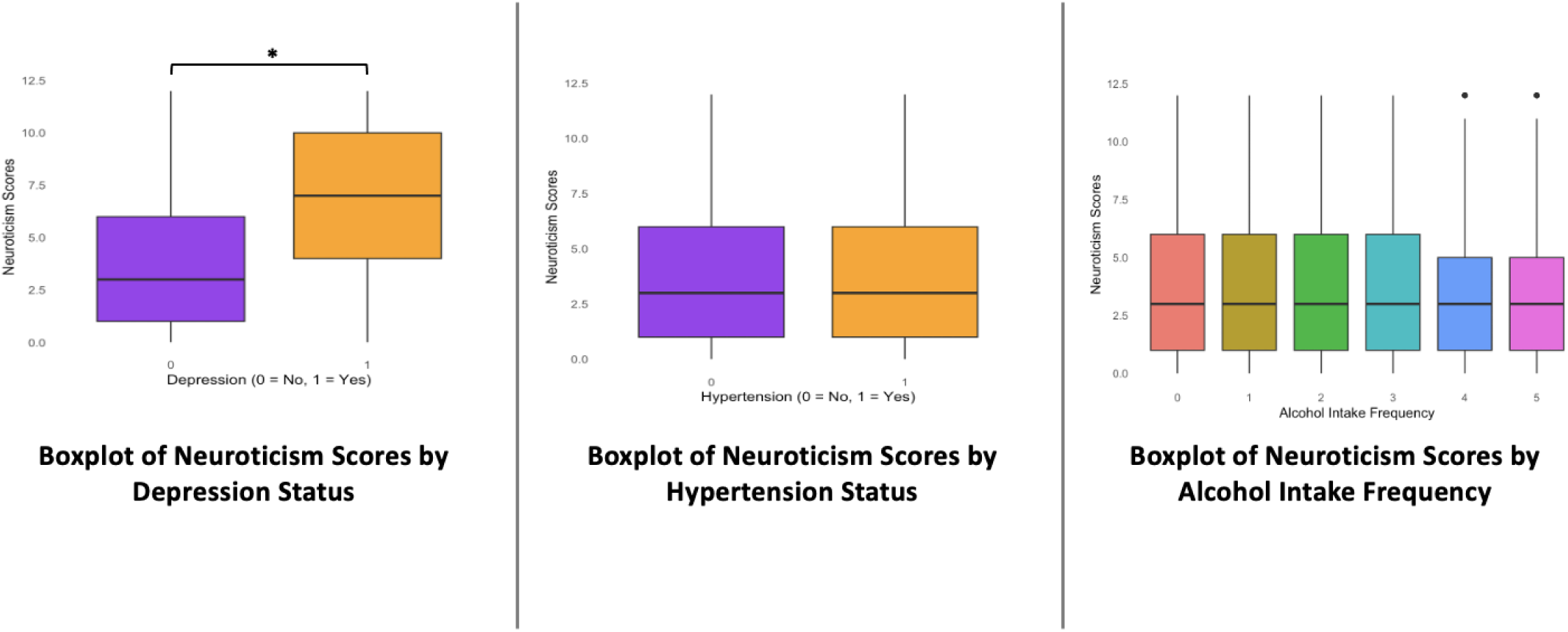
The Difference in Neuroticism Scores Among Participants with and without Depression, with and without Hypertension, and with Different Alcohol Intake Frequencies. Boxplots display the distribution of neuroticism scores stratified by (left) depression status (0=No, 1=Yes), (middle) hypertension status (0=No, 1=Yes), and (right) alcohol intake frequency (0=never, 1=special occasions only, 2=once or three times a month, 3=once or twice a week, 4=three or four times a week, 5=daily or almost daily). Only a significant difference (*p*<0.05) was observed in neuroticism scores between participants with and without a life-time history of depression, indicated by the asterisk.

Last, to explore whether the shift to negative association was influenced by collinearity among the significant mediators and neuroticism, correlations among these variables were assessed. As demonstrated in Figure 6, individuals with depression exhibited significantly higher neuroticism scores, indicating a potential collinearity between neuroticism and depression.

## 4. Discussion

Recent epidemiological studies (see introduction) have identified the personality trait of neuroticism as a potential risk factor for AD. Although the mechanisms underlying the association between neuroticism and AD remain unclear, several models have been proposed to explain this relationship. One possibility is that neuroticism may directly increase the risk of AD. Alternatively, it may exert its influence indirectly by affecting modifiable health and behavioral factors, including hypertension, depression, smoking, alcohol consumption, BMI, and educational attainment, that are themselves established risk factors for AD (Segerstrom, 2020). To our knowledge, this is the first study to investigate both the potential causal relationship between neuroticism and AD, as well as the mediating role of these modifiable factors.

First, the small effect sizes and inconsistent statistical significance across different MR models and GWAS for neuroticism and AD weaken the evidence for a causal relationship. However, we identified Hypertension (β=0.005, p=2.1×10⁻⁴), depression (β=0.048, p=3.3×10⁻⁴), and alcohol consumption (β=0.001, p=1×10⁻⁵) as significant mediators of the neuroticism-AD association, with depression showing the strongest influence, followed by hypertension and alcohol. In other words, a history of depression and hypertension, along with increased alcohol use, may partially explain the relationship between higher levels of neuroticism and increased risk of AD. These findings are consistent with previous research showing that neuroticism is associated with an increased risk of hypertension (Lone and Othman Albotuaiba, 2023; Zhang et al., 2021), depression (Kotov et al., 2010; Speed et al., 2019), and increased alcohol use (Adan et al., 2017; Turiano et al., 2012), which are, in turn, risk factors for AD (Livingston et al., 2020).

Indeed, heightened activation of the sympathetic nervous system and hypothalamic-pituitary-adrenal (HPA) axis in individuals with high neuroticism offers a plausible biological mechanism underlying the mediating relationships observed in this study. Individuals with high levels of neuroticism tend to perceive ordinary situations as threatening, experience negative emotions more intensely and frequently, and overreact to many sources of stress (Barlow et al., 2013). This chronic emotional instability and stress exposure can lead to overactivation of the sympathetic nervous system and HPA axis, along with dysregulation of immune and inflammatory responses, which, in turn, have a direct physiologic effect on the cardiovascular system, resulting in hypertension and development of coronary heart disease (Ayada et al., 2015; Spruill, 2010). Furthermore, the dysregulation of the HPA axis may also result in altered cortisol levels (Montoliu et al., 2020; Tyrka et al., 2006) ,which can impair the body’s ability to manage stress and maintain emotional balance, increasing the risk of depression and mood disorders (Mikulska et al., 2021). It has also been shown that individuals with high levels of neuroticism are more likely to use alcohol to alleviate their distressing emotions (Woicik et al., 2009), which may further exacerbate the risk of adverse health outcomes. Thus, hypertension, depression, and increased alcohol use increase the risk of AD by affecting multiple neurobiological pathways. Hypertension contributes to alterations in the microvascular environment of the CNS, leading to endothelial dysfunction, reduced cerebral blood flow, regional hypoxia, and metabolic abnormalities (Sierra, 2020; Sáiz-Vazquez et al., 2022), which may increase beta-secretase 1 activity and promote amyloid plaque formation, a hallmark of AD pathology. Depression can result in excessive corticosteroid exposure in the hippocampus, which can damage hippocampal neurons, reduce hippocampal volume, and impair neurogenesis in the dentate gyrus (Cereseto et al., 2006; Czéh & Lucassen, 2007; Leuner & Gould, 2010), thereby decreasing memory and increasing dementia risk. Chronic alcohol use further exacerbates AD risk through excitotoxicity and oxidative stress, alterations to gut microbiota, and increased intestinal permeability, which trigger systemic inflammation and neuroinflammation. These processes enhance the neurotoxicity of the amyloid-beta cascade (Bishehsari et al., 2017; Peng et al., 2020).

It is also important to consider that personality traits, including neuroticism, may change during the progression of dementia. Longitudinal studies suggest that individuals may experience increases in neuroticism and decreases in traits such as conscientiousness and openness as neurodegeneration advances, particularly affecting frontal and temporal brain regions (Yoneda et al., 2017). These dynamic changes highlight the need to interpret baseline personality measures cautiously, as the observed associations with AD risk may differ from associations later in the disease course.

Another finding of this study was that when controlling for identified mediators, the direct association between neuroticism and AD became negative. The reversal in the direction of the direct neuroticism–AD association after adjustment for mediators, particularly depression, is likely attributable to statistical suppression arising from strong collinearity between neuroticism and depression. Thus, this finding should be interpreted cautiously as a statistical artifact rather than definitive evidence of a protective effect. Nevertheless, consistent with prior work, it remains possible that certain aspects of neuroticism, when disentangled from comorbid psychopathology, may be associated with positive health outcomes (Friedman, 2000, 2019; Akbarian, 2024). For instance, higher neuroticism has been associated with an 8% reduction in all-cause mortality (HR=0.92, 95% CI=[0.89, 0.95]) after adjusting for demographic, behavioral, and clinical factors (Gale et al., 2017). Prior findings also suggest that neurotic individuals may be more health-conscious and proactive in seeking medical care (Cuijpers et al., 2010; Weston & Jackson, 2018), potentially increasing engagement with preventive interventions that reduce disease risk.

This study had several strengths, including the use of MR, which strengthens causal inference by minimizing confounding and reverse causation. It also leveraged a large sample of healthy controls and older adults diagnosed with AD, along with comprehensive data on health and behavioral factors for mediation analysis. However, the study’s results need to be viewed in light of their limitations. First, in MR analysis, although genome-wide significant SNPs were used as instrumental variables, the genetic instruments for neuroticism account for only a modest proportion of phenotypic variance, raising the possibility of weak-instrument bias and reduced statistical power. Second, horizontal pleiotropy represents a major challenge for MR analyses of psychiatric and behavioral traits, which are highly polygenic and influenced by variants with pleiotropic effects. While multiple sensitivity analyses were performed, including MR-Egger, weighted median, MR-Radial, and MR-PRESSO, residual pleiotropy and violations of MR assumptions cannot be fully excluded. In addition, significant heterogeneity across SNPs was observed in several models, suggesting that not all instruments may fully satisfy MR assumptions. Although leave-one-out analyses and outlier removal did not materially alter the direction of effect, these findings underscore the need for cautious interpretation. Finally, the use of GWAS data predominantly derived from individuals of European ancestry limits the generalizability of these results to other populations. Accordingly, the MR results should be viewed as supportive rather than definitive evidence against a causal relationship between neuroticism and AD. Furthermore, in the mediation analysis, diagnoses for AD, hypertension, and depression were based on survey responses and medical records, which may lack clinical depth and diagnostic accuracy. Moreover, the analyses did not account for the timing of diagnoses for health conditions including AD or model time to dementia, which limits inference about temporal relationships and comparability with survival-based analyses. Nevertheless, the findings of this study suggest that the relationship between neuroticism and AD is primarily mediated by factors such as depression, hypertension, and alcohol consumption, rather than representing a direct causal effect. Future studies should investigate additional potential mediators of the neuroticism-AD relationship to gain a more comprehensive understanding of the underlying mechanisms to inform targeted intervention strategies.

## Data Availability

All data produced in the present study are available upon reasonable request to the authors.

## Acknowledgments

We would like to thank Marcos Sanchez for his invaluable insights into our statistical analyses. We also express our sincere gratitude to all the participants of the UK Biobank, as well as Judy and Larry Tanenbaum Family Foundation, Canadian Institutes of Health Research Fellowship, and Krembil Foundation.

